# Vitamin D receptor polymorphisms in overweight/obese chronic kidney disease patients undergoing hemodialysis

**DOI:** 10.1101/2021.06.02.21258095

**Authors:** Gokhan Bagci, Can Huzmeli, Ferhan Candan

## Abstract

**Background:** Many studies were carried out to investigate the relationship between single nucleotide polymorphisms (SNPs) in vitamin D receptor (VDR) gene with obesity. However, little is known about the role of VDR gene polymorphism with obesity in hemodialysis (HD) patients. Therefore, we aimed to investigate VDR gene TaqI, ApaI and FokI SNPs in overweight/obese HD patients.

**Methods:** Seventy one normal weight and 68 overweight/obese HD patients were included in study. PCR-RFLP method was used for genotyping. Demographic and laboratory data obtained from medical records of patients.

**Results:** For all three SNPs, no significant association was found between normal and overweight/obese patients (P>0.05). Lower HDL concentrations and higher levels of triglyceride (TG) and glucose were detected in the obese/overweight patients compared to normal weight (p<0.001 for HDL, and TG and p=0.023 for glucose). In obese/overweight patients, subjects with CC genotype of TaqI showed higher PTH level (717.1±616.4 pg/ml) than those TC genotype (342.7±360.8 pg/ml) and TT genotype (310.2±323.4 pg/ml) (p=0.028); higher TG level was found in patients with CC genotype of ApaI (627.3±653.0 mg/dl) compared to AA (223.3±156.6) and AC genotypes (193.1±85.4) (p<0.001). Obese/overweight patients carrying FokI TT genotype had higher glucose concentration compared to those carrying CC and CT genotypes (CC=183.4±128.4 mg/dl; TT=151.9±66.1 mg/dl; CT=107.6±41.9 mg/dl, p=0.008).

**Conclusions:** Our study suggest that VDR TaqI, ApaI and FokI polymorphisms are not associated with obesity in HD patients. However, they might be increase the risk of secondary hyperparathyroidism, dyslipidemia, and hyperglycemia, which are among the most common obesity related comorbidities of chronic kidney disease.

## Introduction

Obesity is a chronic multifactorial disease in which genetic and environmental factors play a role. Obesity is associated with important diseases such as type 2 diabetes, cardiovascular disease (CVD), hypertension and cancer (1). It is a major public health problem affecting an important proportion of the world population. According to TURDEP-II study finished in 2010 (26,499 people were participated), the prevalence of obesity in Turkey is 36% and the prevalence of overweight is 37% (2). Obesity is a major risk factor for chronic kidney disease (CKD) and is a strong risk factor for the loss of kidney function after dialysis therapy is started (3).

Vitamin D or 1,25-dihydroxyvitamin D 1,25(OH)2D is a steroid pro-hormone mostly synthesized from UVB-induced 7-dehydrocholesterol in the skin. The fat-soluble cholecalciferol and ergocalciferol are metabolized to a more polar form, 25(OH)D, by the liver. 25(OH)D is hydroxylated to the biologically active form 1,25(OH)2D with renal 1-alpha hydroxylase. Due to a longer half-life and a 1000-fold higher serum concentration, 25(OH)D concentrations are predominantly used to indicate vitamin D deficiency (4).

Vitamin D receptor (VDR) is a nuclear receptor that binds to 1,25(OH)2D with high affinity. VDR forms a heterodimer with the retinoid X receptor and then binds to hormone response elements in the target genes (5). The effect of 1,25(OH)2D is mediated by VDR, which regulates the transcription of many target genes involved in various physiological processes (6, 7). VDR expression was detected in the majority of human tissues, including osteoblasts, smooth muscle cells, macrophages, epithelial cells and pancreatic β-cells, and its expression also highly found in adipocytes. Adipose tissue express both the VDR and the enzymes involved in vitamin D metabolism. VDR expression is reported in human subcutaneous adipose tissue and visceral adipose tissue (8-10).

Vitamin D deficiency is common in CKD patients. Various factors, including increased age and comorbid conditions, such as diabetes and hypertension, have been associated with lower 25(OH)D levels in individuals with both dialysis-dependent and non-dialysis-dependent CKD (11). The main reason for progressive reduction of active vitamin D serum levels in CKD is the simultaneous decrease in renal 1α-hydroxylase levels with progressive decrease in renal mass and function. Furthermore, secondary hyperparathyroidism and hyperphosphoremia may contribute to the inhibition of vitamin D activation by the kidneys. Because of the complex role of vitamin D in CKD, some variants in the VDR gene have been considered as a way of improving the management of the disease since they are thought to affect response to therapeutic approaches (12-13). It is thought that VDR gene polymorphisms may play a role in regulating adipose tissue activity, body fatness and susceptibility to adiposity (10).

Several studies have been performed to investigate VDR polymorphism in CKD patients with comorbidities such as secondary hyperparathyroidism (13), left ventricular hypertrophy (14) atherosclerosis (15), and type II diabetes (16). However, we did not find any study investigating the relationship between obesity and VDR gene polymorphisms in CKD patients. In the light of this information, we aimed to evaluate VDR gene polymorphisms in obese/overweight and non-obese/overweight CKD patients undergoing HD. A secondary objective was to investigate whether there is a relationship between VDR polymorphisms and demographic and biochemical data in obese/overweight HD patients.

## Materials and methods

### Subjects

For this cross-sectional study, we included a total of 139 HD patients, who have applied to the clinic in Department of Nephrology, Cumhuriyet University, and monitored in HD centers in Sivas City or its districts, were screened. According to body mass index (BMI) classification, patients with a BMI ≥25 and <30 were assessed as having overweight, and ≥30 assessed as having obese. Patients with a BMI ≥18 and < 25 were assessed as having normal weight. According to this classification, 68 patients were considered obese or overweight, and 71 patients were considered normal weight. Demographic characteristics and routinely available laboratory data were extracted from the patients’ medical files and from the electronic database of hospital. Patients with a BMI less than 18, patients with active infection or malignancy were excluded from the study.

Fasting blood specimens were collected to measure blood chemistry parameters. 25(OH)D, total cholesterol, triglyceride (TG), high-density lipoprotein (HDL), low-density lipoprotein (LDL), C-reactive protein (CRP), creatinine, calcium, phosphorus, potassium, sodium, blood urea nitrogen (BUN), PTH, protein, albumin, glucose concentrations were measured by routine laboratory methods. Cholesterol to HDL ratio, TG to HDL ratio, and LDL to HDL ratio were calculated.

### Genotyping

After sampling 5 ml of blood from the patients with K3DTA tubes, the samples were stored at -20 °C. The sampling was done using Genomic DNA QIAamp DNA Blood Mini Kit (QIAGEN, Germany).

Polymerase Chain Reaction-Restriction Fragment Length Polymorphism (PCR-RFLP) method was used for genotyping. Primers used for PCR reaction were as follows: TaqI (rs731236) F:5′-CAG AGC ATG GAC AGG GAG CAA G-3′ and R: 5′-GCA ACTC CTC ATG GCT GAG GTC TCA-3′, ApaI (rs7975232) F: 5′ CAG AGC ATG GAC AGG GAG CAA G-3′ and R: 5′-GCA ACT CCT CAT GGC TGA GGT CTC A-3′, and FokI (rs2228570) F: 5′-AGC TGG CCC TGG CAC TGA CTC TTG CTC T-3′ and R: 5′-ATG GAA ACA CCT TGC TTC TTC TCC CTC-3′. The PCR conditions were: denaturation at 94 °C for 15 s, annealing at 63 °C for TaqI and ApaI, and 65 °C for FokI for 30 s, and extension at 72 °C for 30 s for 35 cycles. Final extension at 72 °C for 7 min. PCR products were digested separately with restriction enzymes TaqI, ApaI and FokI (Thermo Fisher Scientific, Waltham, MA, USA). The digested products were analyzed by electrophoresis in a 2% agarose gel stained with ethidium bromide, and then evaluated at UV light.

### Statistical Analysis

All statistical analyses of the study were performed by Statistical Package for the Social Sciences (SPSS) version 22.0 (SPSS IBM, New York, USA). Normally distributed continuous variables were expressed as mean ± standard deviation (SD), and non-normally distributed variables as medians and interquartile ranges (IQR; 25-75th percentiles). Student t-test or Mann-Whitney U test was used for group comparisons. Chi-Square test or Fisher’s exact test was used for the evaluation of categorical values including genotype and allele frequencies of VDR SNPs. One-way ANOVA test was used to compare selected parameters in obese/overweight patients according to genotypes of TaqI, ApaI and FokI SNPs. Statistical significance was defined as P value less than 0.05. Results were expressed with a 95% confidence interval.

## Results

In the present study, we evaluated VDR gene polymorphisms in 68 obese/overweight and 71 normal weight CKD patients undergoing HD and we investigated whether there is a relationship between VDR gene polymorphisms and demographic and biochemical data in obese/overweight HD patients. Demographic and biochemical data of patients are shown in Table 1. As expected, height, weight and BMI was significantly different in the two groups (p = 0.028 for height, p < 0.001 for weight and BMI). The frequency of women in obese/overweight patients was higher than normal weight patients (67.6% to 43.7%, p = 0.004). 52.9% of the patients in the obese/overweight group were diabetes mellitus and 22.5% of the patients in the normal weight group were diabetes mellitus. Lower HDL concentrations and higher levels of TG and glucose were detected in the obese/overweight patients (p<0.001 for HDL and TG and p = 0.023 or glucose). There were no statistically significant differences between the groups in terms of other demographic parameters and laboratory findings.

**Table 1.** Baseline characteristics of obese/overweight and non-obese/overweight hemodialysis patients

| Characteristic | Overall<br>(N = 139) | Obese / overweight<br>(N = 68) | Normal weight<br>(N = 71) | P |
| --- | --- | --- | --- | --- |
| Age (years) | 58.6 ± 14.1 | 60.5 ± 10.3 | 56.8 ± 16.8 | 0.125 |
| Sex (M/F) % | 44.6 / 55.4 | 32.4 / 67.6 | 56.3 / 43.7 | **0.004** |
| Height (cm) | 165 (160-170) | 163 (159-167) | 168 (160-170) | **0.028** |
| Weight (kg) | 68.8 ± 13.2 | 78.3 ± 11.1 | 59.6 ± 7.2 | **<0.001** |
| BMI (kg/m<sup>2</sup>) | 24.8 (21.3-28.3) | 28.4 (26.8-31.5) | 21.5 (20.4-23.5) | **<0.001** |
| Smoking (%) | 15 (10.8) | 5 (9.1) | 10 (16.7) | 0.177 |
| Duration of dialysis (months) | 48 (27-96) | 54 (24-94) | 42 (27-96) | 0.657 |
| Diabetes mellitus (n, %) | 52 (37.4) | 36 (52.9) | 16 (22.5) | **<0.001** |
| Hypertension (n, %) | 25 (18) | 13 (19.1) | 12 (16.9) | 0.733 |
| 25(OH)D (ng/mL) | 10.4 (7.9-13.7) | 10.3 (7.8-14.9) | 10.8 (8.1-12.9) | 0.757 |
| Cholesterol (mg/dL) | 165 (139-206) | 183 (139-213) | 160 (139-200) | 0.114 |
| TG (mg/dL) | 156 (109-239) | 191 (133-274) | 133 (87-184) | **<0.001** |
| HDL (mg/dL) | 34 (27-40) | 31 (25-37) | 37 (29-46) | **<0.001** |
| LDL (mg/dL) | 99 (73-136) | 110.5 (77-145) | 96 (72-125) | 0.112 |
| CRP (mg/L) | 9.8 (2.8-29) | 10.5 (3.9-40) | 6.7 (2.1-21) | 0.119 |
| Creatinine (mg/dL) | 7.2 (6.0-9.6) | 7.5 (6.3-9.7) | 6.9 (5.6-9.6) | 0.454 |
| Calcium (mg/dL) | 8.7 (8.2-9.1) | 8.6 (8.3-9.1) | 8.7 (8.2-9.2) | 0.939 |
| Phosphorus (mg/dL) | 4.4 (3.6-5.4) | 4.6 (3.7-5.4) | 4.2 (3.6-5.5) | 0.499 |
| Potassium (mEq/L) | 5.2 (4.4-5.7) | 5.2 (4.5-5.7) | 5.1 (4.4-5.9) | 0.938 |
| Sodium (mEq/L) | 138 (135-140) | 138 (137-140) | 138 (136-140) | 0.314 |
| BUN (mg/dL) | 64.5 ± 18.3 | 63.7 ± 16.8 | 65.2 ± 19.8 | 0.647 |
| PTH (pg/ml) | 255 (120-488) | 238 (127-470) | 262 (109-513) | 0.583 |
| Protein (g/dL) | 6.8 (6.5-7.2) | 6.9 (6.5-7.3) | 6.8 (6.4-7.0) | 0.066 |
| Albumin (g/dL) | 3.9 (3.6-4.2) | 3.8 (3.6-4.2) | 3.9 (3.7-4.2) | 0.299 |
| Glucose (mg/dL) | 105 (86-147) | 116 (90-155) | 100 (84-121) | **0.023** |
| EPO use (n, %) | 92 (66.2) | 46 (67.6) | 46 (64.8) | 0.858 |
| Calcitriol use (n, %) | 49 (35.3) | 24 (35.3) | 25 (35.2) | 0.992 |
| Calcium use (n, %) | 85 (61.2) | 42 (61.8) | 43 (60.6) | 0.885 |

Data were expressed as mean ± SD, median and interquartile range, or as frequency and percent, as appropriate.

Table 2 show that genotype and allele frequencies of VDR gene SNPs in obese/overweight and normal weight patients. For all three SNPs, no significant difference was found between normal and overweight/obese patients (p > 0.05). Table 3 demonstrates that comparisons of some selected parameters according to TaqI, ApaI and FokI genotypes of VDR gene in overweight/obese patients. The genotypes of TaqI SNP of VDR gene did not show any significant differences in all selected parameters except from PTH level. The CC genotype of the TaqI VDR showed higher PTH level (717.1±616.4 pg/ml) than the TC genotype (342.7±360.8 pg/ml) and the TT genotype (310.2±323.4 pg/ml) (p = 0.028). The genotypes of ApaI-VDR did not show any statistically significant differences in all selected parameters except from TG level and TG to HDL ratio. Higher TG level was found in the CC genotype of the ApaI-VDR (627.3 ± 653.0 mg/dl) compared to AA (223.3±156.6) and AC genotypes (193.1±85.4) (p < 0.001). Similarly, higher TG to HDL ratio was found in the ApaI CC genotype (25.3±26.1) compared to the AA genotype (8.4±7.1) and the AC genotype (7.1±4.8). There were no significant differences in the selected parameters among the three genotypes of the FokI SNP except for glucose level. Obese/overweight patients carrying FokI TT genotype had higher glucose concentration compared to CC and CT genotypes (CC: 183.4±128.4; TT: 151.9±66.1; CT: 107.6±41.9, p = 0.008).

**Table 2.**
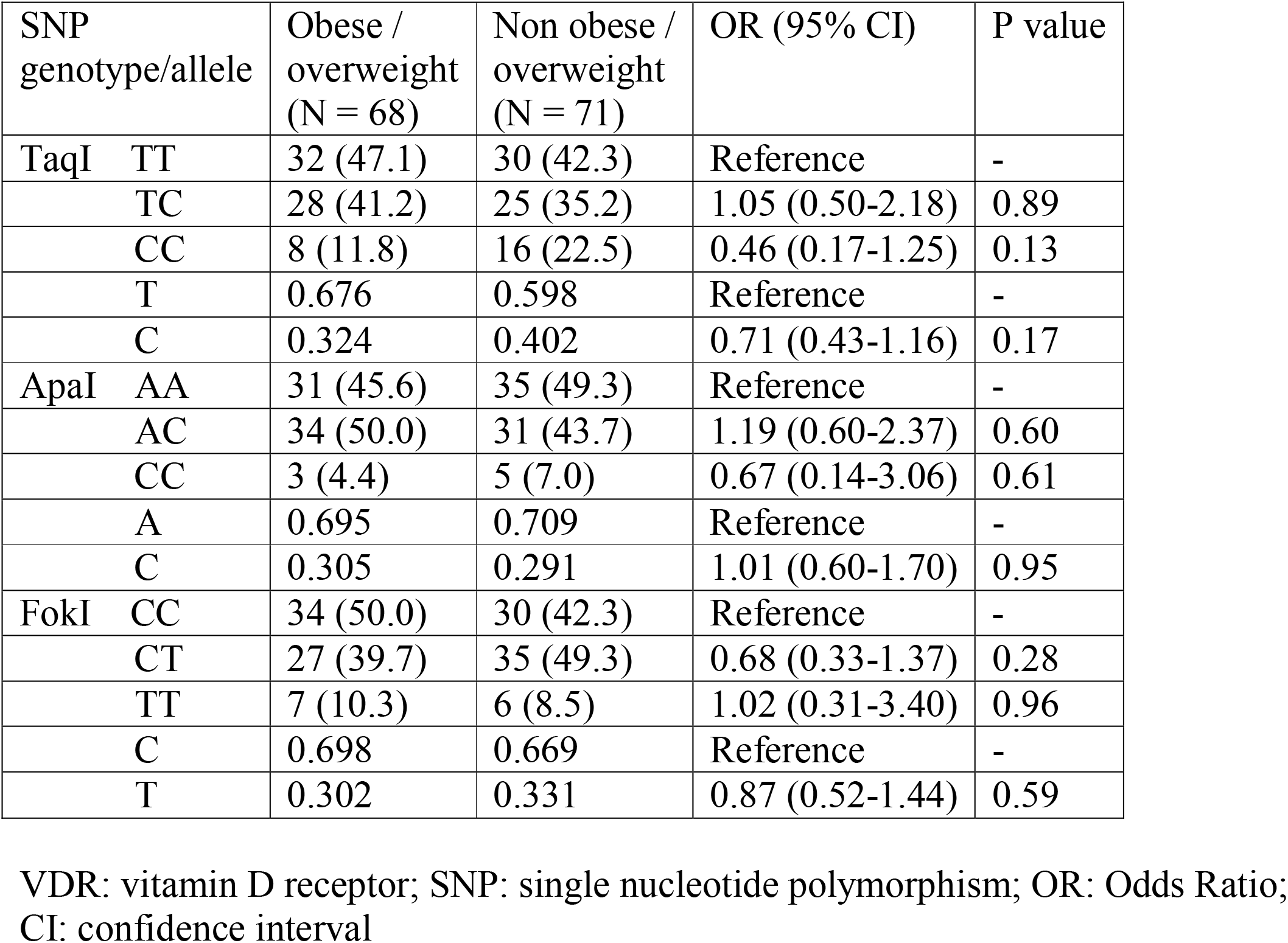
Genotype and allele frequencies of VDR gene SNPs in obese/overweight and normal weight hemodialysis patients

**Table 3.** Comparisons of some selected parameters according to VDR gene TaqI, ApaI and FokI genotypes in obese/overweight patients.

|  | TaqI |  |  |  | ApaI |  |  |  | FokI |  |  |  |
| --- | --- | --- | --- | --- | --- | --- | --- | --- | --- | --- | --- | --- |
| Parameter | TT | TC | CC | P | AA | AC | CC | P | CC | CT | TT | P |
| BMI (kg/m<sup>2</sup>) | 29.1(3.4) | 30.0(4.6) | 29.7(4.1) | .70 | 29.3(3.4) | 29.9(4.6) | 27.8(1.8) | .62 | 29.4(4.5) | 29.5(3.6) | 30.6(3.2) | .74 |
| 25(OH)D (ng/ml) | 11.3(4.3) | 11.4(6.0) | 11.8(5.0) | .47 | 10.9(3.9) | 11.4(4.5) | 16.8(4.4) | .08 | 11.3(4.2) | 11.7(4.5) | 11.3(4.9) | .93 |
| Cholesterol (mg/dl) | 176.9(55.1) | 181.3(44.2) | 154.6(53.4) | .17 | 182.6(56.2) | 187.3(64.8) | 137.7(33.0) | .39 | 188.6(61.7) | 174.9(63.8) | 185.7(39.0) | .69 |
| TG (mg/d) | 210.2(195.8) | 183.4(103.8) | 166.9(86.8) | .81 | 223.3(156.6) | 193.1(85.4) | 627.3(653.0) | **<.001** | 224.4(138.6) | 246.7(252.9) | 198.1(78.3) | .80 |
| HDL (mg/dl) | 33.9(10.2) | 36.7(14.0) | 34.8(12.4) | .67 | 32.3(10.4) | 31.3(9.1) | 25.3(3.5) | .48 | 31.7(10.3) | 30.7(8.4) | 33.1(11.0) | .82 |
| LDL (mg/dl) | 138.0(137.6) | 112.5(48.7) | 108.1(38.4) | .56 | 134.6(137.3) | 116.7(51.9) | 96.7(65.2) | .69 | 142.0(130.5) | 103.0(54.5) | 117.7(37.7) | .32 |
| Cholesterol/HDL | 6.0(1.9) | 6.5(2.1) | 5.2(2.3) | .28 | 6.0(2.3) | 6.2(2.0) | 5.4(1.1) | .76 | 6.4(2.5) | 5.7(1.6) | 5.9(1.3) | .42 |
| TG/HDL | 8.9(10.2) | 8.4(6.7) | 7.3(3.2) | .88 | 8.4(7.1) | 7.1(4.8) | 25.3(26.1) | **.001** | 8.4(6.5) | 9.0(10.8) | 7.0(4.4) | .84 |
| LDL/HDL | 4.6(5.9) | 3.6(1.3) | 3.8(1.0) | .62 | 4.5(6.0) | 3.8(1.2) | 3.7(2.0) | .74 | 3.0(1.4) | 2.9(1.3) | 2.1(0.7) | .31 |
| Creatinine (mg/dl) | 7.8(2.5) | 8.0(2.9) | 8.4(1.6) | .83 | 7.8(2.6) | 8.1(2.6) | 7.3(2.8) | .79 | 7.4(2.5) | 8.4(2.5) | 8.5(2.8) | .29 |
| Calcium (mg/dl) | 8.7(0.8) | 8.5(0.7) | 8.6(0.5) | .52 | 8.7(0.8) | 8.5(0.7) | 9.3(0.8) | .18 | 8.4(0.9) | 8.8(0.8) | 8.6(1.3) | .85 |
| Phosphorus (mg/dl) | 4.8(1.3) | 4.3(1.4) | 4.8(1.1) | .34 | 4.6(1.3) | 4.6(1.4) | 4.3(1.2) | .90 | 4.6(1.2) | 4.6(1.6) | 4.3(1.0) | .79 |
| BUN (mg/dl) | 67.0(14.0) | 61.7(19.0) | 58.1(18.4) | .29 | 67.0(18.2) | 60.8(15.6) | 64(14.8) | .34 | 60.3(15.9) | 65.4(17.0) | 74.3(17.2) | .11 |
| PTH (pg/ml) | 310.2(323.4) | 342.7(360.8) | 717.1(616.4) | **.028** | 480.0(481.0) | 286.2(298.3) | 261.7(22.7) | .13 | 258.9(221.3) | 489.3(534.7) | 484.7(350.6) | .06 |
| Glucose (mg/dl) | 133.8(54.9) | 138.6(78.8) | 148.8(101.5) | .86 | 133.2(83.8) | 139.2(60.0) | 163.7(33.6) | .76 | 151.9(66.1) | 107.6(41.9) | 183.4(128.4) | **.008** |

## Discussion

Adipose tissue is an organ which has important metabolic functions. It plays an important role in energy balance and glucose hemostasis. It is also known as the main storage source for vitamin D (17). Obesity is caused by the imbalance between energy intake and expenditure. Obese people have increased body fat mass (adipocyte hyperplasia and hypertrophy). Obesity, an outbreak of the 21st century, is an increased risk factor for comorbid complications such as CVD, hypertension, dyslipidemia, type 2 diabetes and cancer. In addition, obesity increases the risk of CKD and its progression to end stage renal disease (18). Obesity and diabetes mellitus are very common in HD patients. It has been suggested that an association was found between vitamin D deficiency and obesity (3).

The relationship between vitamin D deficiency and obesity is explained in various ways: i) Obese individuals have reduced exposure to the sun compared to the weak, ii) Vitamin D is sequestered in fat tissue, iii) Low 1.25(OH)2D inhibits adipogenesis, iiii) Low 25(OH)D concentration is just due to volumetric dilution (17-19). It has been suggested that vitamin D is associated with dyslipidemia (20). The administration of 1,25(OH)D significantly increases the calcium (Ca^+2^) transport into the adipose tissue and thereafter lipogenesis is induced and lipolysis is reduced (21). In addition, 1,25(OH)2D can stop the production of uncoupling protein (UCP2) by accelerating the Ca^+2^ flow into the cell, and thus it has the effect of increasing obesity (22).

In Turkey, vitamin D deficiency and insufficiency have been commonly detected in overweight and obese individuals, especially in women and there was a negative correlation between vitamin D level and obesity (24). González et al. (25) found significantly lower vitamin D levels in individuals with high BMI, waist circumference (WC), and waist-to-height ratio. Saneei et al. (26) included 34 studies in the meta-analysis, they reported that a significant inverse weak correlation between BMI and serum 25(OH)D levels. Pereira-Santos et al. (27) included 23 studies in the meta-analysis, they found that obese subjects had 35% vitamin D deficiency compared to those with normal weight and 24% higher than in the overweight group. In another meta-analysis. Yao et al. (28) reported that vitamin D deficiency was prevalently associated with obesity in European-Americans and Asians. The prevalence of vitamin D deficiency is higher among the CKD, diabetes and obesity patients in the groups which are major risk factors for vitamin D deficiency (29).

VDR is present on adipocytes and many studies have been carried out to examine the relationship between VDR gene polymorphisms and obesity. In a study with Pakistani patients, no significant relationship was found between VDR FokI SNP and obesity (30). In a study with obese women, vitamin D deficiency and insufficiency were detected in 80.3% of the subjects. No significant difference was found between the two groups in terms of Apa1 polymorphism. (31). In Chinese population, the TT genotype and the T allele of TaqI SNP is found to be associated with obesity (32). A study in the Greek population showed that TaqI T allele associated with higher BMI (33). In another study, ApaI and TaqI SNP of the VDR gene were examined in patients with diabetes mellitus. No significant difference has been shown between patients and controls. However, in subjects with early-onset T2DM, a higher BMI and an increased prevalence of obesity (81%) was found in subjects carrying TT genotype of the TaqI compared to TC (46%) or tt (52%) genotypes (34). In the 1958 British birth cohort study (1958BC) which was conducted with 5,224 participants, VDR SNPs were not associated with obesity traits (35). In a cross-sectional study of 198 Arab adults, the TT carriers of FokI SNP had higher total cholesterol than that of CC and CT genotypes (36). In a recent study conducted with 402 obese and 489 non-obese Saudis, VDR TaqI minor allele polymorphisms were found more frequent in obese individuals (37).

In the current study, lower HDL and higher TG concentrations was detected in obese/overweight HD patients compared to non-obese/overweight patients. In a study conducted in obese children, a positive correlation was found between serum 25(OH)D concentration and HDL, while a negative correlation was found between TG (14). In a study of 1534 individuals, a positive correlation was found between vitamin D, HDL and age. A negative correlation was found between vitamin D and BMI, LDL, cholesterol and TG (38). NHANES III trial demonstrated negative association between serum 25(OH)D levels and hypertriglyceridemia, diabetes, hypertension, and obesity (39). In children with obesity and metabolic syndrome, vitamin D supplementation did not affect total cholesterol, LDL and HDL levels, but TG levels decreased (40). Jorde et al. (41) included 12 cross-sectional and 10 placebo-controlled intervention studies in the meta-analysis, they found a positive correlation between serum 25(OH)D level and HDL and a negative correlation with TG in the majority of studies. On the other hand, recent randomized clinical trials evaluating the effect of vitamin D supplementation on blood lipids have reported conflicting evidences. In the majority of studies, it has been found that vitamin D supplementation is not a significant effect on blood lipids compared to placebo (42-44). These recent findings make it difficult to decide whether there is a causal relationship between vitamin D deficiency and a negative blood lipid profile.

In the present study, glucose concentration was higher in the obese/overweight group. This may be due to the high number of patients with diabetes mellitus in the obese/overweight group. 1,25(OH)2D is an important regulator of insulin release from pancreas (45). In 1,25(OH)2D deficient animals, intravenous 1.25(OH)2D administration improves glucose intolerance and induces insulin releasing from beta cells of pancreas in response to a glucose challenge (46). In a study involving 120 non-diabetic CKD patients, obese patients treated with 1,25(OH)2D had similar insulin concentrations compared to non-obese patients, whereas untreated obese patients had a higher insulin concentration (47). In our study, obese/overweight patients carrying FokI homozygous mutant TT genotype had higher glucose concentrations compared to those having CC and CT genotypes. In another study, individuals with FokI TT genotype was associated with higher HOMA-IR values than individuals with CT genotype. Thus, VDR FokI polymorphism has suggested to be associated with diabetes mellitus (48).

It has been suggested that there was a potential association between parathyroid function and VDR polymorphisms in CKD patients (13). In the current study, significantly higher PTH levels were found in obese/overweight patients carrying TaqI CC genotype compared to those carrying TT and TC genotypes. In Iranian HD patients, similar findings were found in terms of TaqI variants (49). It has been suggested that the TaqI TT genotype increases the risk of development of hyperparathyroidism in Turkish HD patients (13).

The main limitation of the current study is the relatively small sample size. Further, we only calculated BMI for evaluation of obesity. We did not measured waist to hip ratio and waist circumference for evaluation of obesity.

In conclusion, our study shows that VDR gene TaqI, ApaI and FokI genotype and allele frequencies are not different in obese/overweight HD patients compared to normal weight patients. However, in obese/overweight HD patients, individuals carrying TaqI CC genotype have higher PTH levels, those carrying ApaI CC genotype have higher TG levels and those carrying FokI TT genotype have higher glucose levels. Although this study has shown that VDR polymorphisms are not related to obesity susceptibility, in overweight/obese HD patients, VDR polymorphisms have been suggested to increase the risk of secondary hyperparathyroidism, dyslipidemia, and hyperglycemia, which are among the most common obesity related comorbidities of CKD. Further studies are required to reveal any possible association between VDR polymorphisms and obesity in HD patients.

## Data Availability

No data

## Compliance with ethical standards

### Ethical approval

Local ethics committee approval was obtained from Clinical Research Ethics Committee of Sivas Cumhuriyet University, Sivas, Turkey. All procedures performed in the current study have been performed in accordance with the ethical standards of the 1964 Helsinki Declaration and subsequent amendments or comparable ethical standards.

### Informed consent

All patients gave a written informed consent to participate in the current study.

